# Sharps Injuries Among Healthcare Workers in Liberia and Ghana: A Cross-Sectional Survey

**DOI:** 10.1101/2023.09.19.23295623

**Authors:** Laura Jean Ridge, John Arko-Mensah, Josh Lambert, Lydia Aziato, G Clinton Zeantoe, Henry Duah, Marjorie McCullagh

## Abstract

**Objective:** There is little data on sharps injuries among healthcare workers in West Africa, despite the region’s high rate of Hepatitis B and HIV. The purpose of this study is to investigate healthcare workers’ history of sharps injury in Liberia and Ghana.

**Design:** An electronic cross-sectional survey disseminated by local nursing, midwifery, physician assistant, and physician associations.

**Setting:** Healthcare workers in Liberia and Ghana from February to June 2022.

**Participants:** Participant were registered nurses, physician assistants, physicians, or midwives, and had been working in a patient care role for at least nine of the previous twelve months.

**Methods:** A link to the survey was texted to participants through their professional association membership lists, including nursing, midwifery, and physician assistant organizations in both Liberia and Ghana and a physician organization in Ghana only.

**Results:** 509 participants reported an average of 1.8 injuries per year in Liberia and 1.1 in Ghana (p=<0.01). 15.1% of healthcare workers reported three or more injuries in the past year. Liberia had a higher proportion of frequently injured workers (p=<0.01). Frequently injured workers were evenly distributed across worker types.

**Conclusions:** Workers in this region are vulnerable to sharps injuries. A frequently injured subset of workers likely has distinctive risk factors and would benefit from further investigation and intervention.

## Introduction

Sharps injuries are a major occupational hazard among healthcare workers worldwide. It is estimated that these injuries cause 16,000 new Hepatitis C infections, 66,000 new Hepatitis B infections, and 1,000 new HIV infections annually (1). Even though reported sharps injury rates are high in low- and middle-income countries (LMICs), surveillance of injury incidence and reporting is low (2,3).

This is particularly true in West Africa (4), despite the enormous occupational health challenges faced by healthcare workers in that region (5), including the highest rate of HBV (∼8%) worldwide (6). It is estimated that mean number of sharps injuries in West Africa per HCW per year is 2.1, resulting in 33,000 exposures to HIV and HCV, and 131,000 exposures to HBV, per year (7). Approximately 41.7% of African healthcare workers reported a sharps injury in the past year, although that data came primarily from other regions in Africa (2).

Liberia, a country of about 5 million people, has about 19,000 healthcare workers, while Ghana, a country of about 31 million people, has about 123,000 healthcare workers (8). Data on sharps injuries in both countries is scant, with only a few facility-specific studies in Ghana (9). One of these studies found that 36.3% of emergency department nurses at a single hospital in Ghana had four or more injuries in the past twelve months, raising the possibility of a larger population of frequently injured workers (10). The purpose of this paper is to investigate the incidence and correlates of sharps injuries among healthcare workers in Liberia and Ghana and to identify subsets or categories of workers with high incidence of injuries.

## Methods

A cross-sectional survey was administered via Qualtrics to healthcare workers in Liberia and Ghana from February to June, 2022. Inclusion criteria included: the participant was a registered nurse, physician assistant, physician, or midwife, and that he or she had been working in a patient care role for at least nine of the previous twelve months. Exclusion criterion was healthcare workers, even from those cadres, who worked primarily as administrators, nursing assistants, or mental health clinicians. A link to the survey was texted to participants through their professional association membership lists including nursing, midwifery, and physician assistant organizations in both Liberia and Ghana, and a physician organization in Ghana only.

The survey instrument was developed by the study team, which included Ghanaian, Liberian, and American researchers with experience in public health and occupational exposures in their respective countries. The survey instrument included questions about demographics and sharps injury experience, both overall and within the past twelve months. A seven-item multiple choice sharps knowledge scale was developed from Liberian and Ghanaian national infection prevention and control policies (11,12). The six-item Safety Culture: Evaluation Survey, developed by the Centers for Disease Control’s (CDC) National Institute for Occupational Safety and Health (13), was also included.

The instrument was reviewed for face and content validity by the leadership of participating local health professional associations, and suggested revisions were made by the PI. The survey link and a brief explanation of the project was texted to professional association membership lists contact lists no more than three times. Liberian participants received $3 USD and Ghanaian participants received 15 Cedis via remote scratch card top-up as compensation.

Descriptive and analytic statistical analysis was conducted in JMP 17 (JMP*®*, Version *17 PRO*. SAS Institute Inc., Cary, NC, 1989–2023) by a trained biostatistician. Continuous variables were analyzed using *t*-tests and ANOVA; categorial variables were tested using Pearson chi-squared test. The STROBE checklist for cross-sectional studies was used to guide reporting within the manuscript (Figure 1).

This project received in-country approval from the University of Liberia’s Institutional Review Board (Protocol #21-12-296) and the Ghana Health Services Ethics Research Committee (GHS ERC Number: 018/11/2021). It was deemed exempt by the Institutional Review Board of the University of Michigan and the University of Cincinnati.

## Results

Nine hundred and seventy-one people were screened for participation; 618 met inclusion criteria. Of those 618 participants, 509 recorded the number of injuries they had in the previous 12 months and were included in the analysis (Table 1). Of the participants, 458 were from Ghana and 51 were from Liberia. All 16 regions in Ghana and 11 of Liberia’s 15 counties were represented in the analysis. Liberian participants were more likely to be female (p<.01) and were older than Ghanaian participants (*p*<.01), although for both groups the largest age group was 31-40 years. Nurses were the largest healthcare worker group represented in both countries.

**Table 1:**
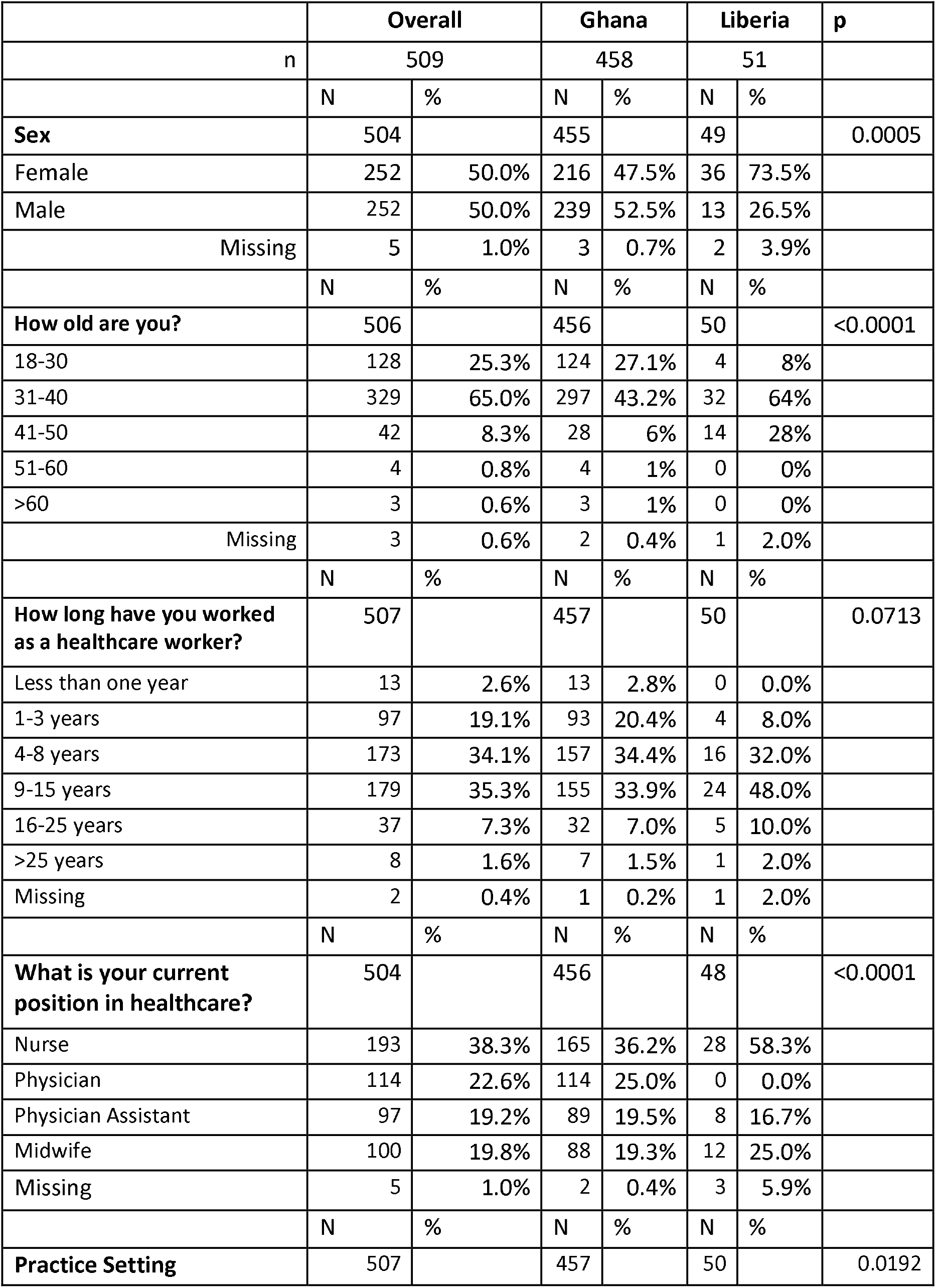
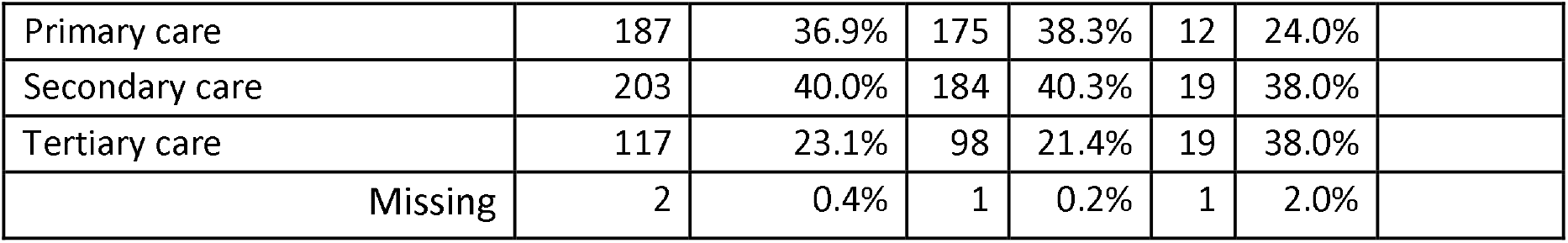
Participants.

|  | Overall |  | Ghana |  | Liberia |  | p |
| --- | --- | --- | --- | --- | --- | --- | --- |
| n | 509 |  | 458 |  | 51 |  |  |
|  | N | % | N | % | N | % |  |
| **Sex** | 504 |  | 455 |  | 49 |  | 0.0005 |
| Female | 252 | 50.0% | 216 | 47.5% | 36 | 73.5% |  |
| Male | 252 | 50.0% | 239 | 52.5% | 13 | 26.5% |  |
| Missing | 5 | 1.0% | 3 | 0.7% | 2 | 3.9% |  |
|  | N | % | N | % | N | % |  |
| **How old are you?** | 506 |  | 456 |  | 50 |  | <0.0001 |
| 18-30 | 128 | 25.3% | 124 | 27.1% | 4 | 8% |  |
| 31-40 | 329 | 65.0% | 297 | 43.2% | 32 | 64% |  |
| 41-50 | 42 | 8.3% | 28 | 6% | 14 | 28% |  |
| 51-60 | 4 | 0.8% | 4 | 1% | 0 | 0% |  |
| >60 | 3 | 0.6% | 3 | 1% | 0 | 0% |  |
| Missing | 3 | 0.6% | 2 | 0.4% | 1 | 2.0% |  |
|  | N | % | N | % | N | % |  |
| **How long have you worked as a healthcare worker?** | 507 |  | 457 |  | 50 |  | 0.0713 |
| Less than one year | 13 | 2.6% | 13 | 2.8% | 0 | 0.0% |  |
| 1-3 years | 97 | 19.1% | 93 | 20.4% | 4 | 8.0% |  |
| 4-8 years | 173 | 34.1% | 157 | 34.4% | 16 | 32.0% |  |
| 9-15 years | 179 | 35.3% | 155 | 33.9% | 24 | 48.0% |  |
| 16-25 years | 37 | 7.3% | 32 | 7.0% | 5 | 10.0% |  |
| >25 years | 8 | 1.6% | 7 | 1.5% | 1 | 2.0% |  |
| Missing | 2 | 0.4% | 1 | 0.2% | 1 | 2.0% |  |
|  | N | % | N | % | N | % |  |
| **What is your current position in healthcare?** | 504 |  | 456 |  | 48 |  | <0.0001 |
| Nurse | 193 | 38.3% | 165 | 36.2% | 28 | 58.3% |  |
| Physician | 114 | 22.6% | 114 | 25.0% | 0 | 0.0% |  |
| Physician Assistant | 97 | 19.2% | 89 | 19.5% | 8 | 16.7% |  |
| Midwife | 100 | 19.8% | 88 | 19.3% | 12 | 25.0% |  |
| Missing | 5 | 1.0% | 2 | 0.4% | 3 | 5.9% |  |
|  | N | % | N | % | N | % |  |
| **Practice Setting** | 507 |  | 457 |  | 50 |  | 0.0192 |

|  |  |  |  |  |  |  |
| --- | --- | --- | --- | --- | --- | --- |
| Primary care | 187 | 36.9% | 175 | 38.3% | 12 | 24.0% |
| Secondary care | 203 | 40.0% | 184 | 40.3% | 19 | 38.0% |
| Tertiary care | 117 | 23.1% | 98 | 21.4% | 19 | 38.0% |
| Missing | 2 | 0.4% | 1 | 0.2% | 1 | 2.0% |

The majority of participants in both countries had between 4 and 15 years of experience as a healthcare worker, with the plurality of Ghanaian healthcare workers reporting 4 to 8 years of work experience, and the plurality of Liberian healthcare workers reporting 9 to 15 (p=0.07). Participants’ work settings were fairly evenly spread across different facility types (primary, secondary, and tertiary); in both countries between 38.0-40.3% of participants worked in secondary care facilities. The remainder of participants worked mostly in primary care facilities in Ghana (40.3%) and tertiary care facilities in Liberia (38.0%) (*p*=0.02).

The mean number of injuries in the past twelve months was 1.1, with 1.8 the average in Liberia and 1.1 the average in Ghana (p=<0.01) (Table 2). The healthcare worker groups with the highest average number of injuries were nurses (1.2) and midwives (1.3), but the difference between the healthcare worker groups was not statistically significant (*p*=0.57).

**Table 2:**
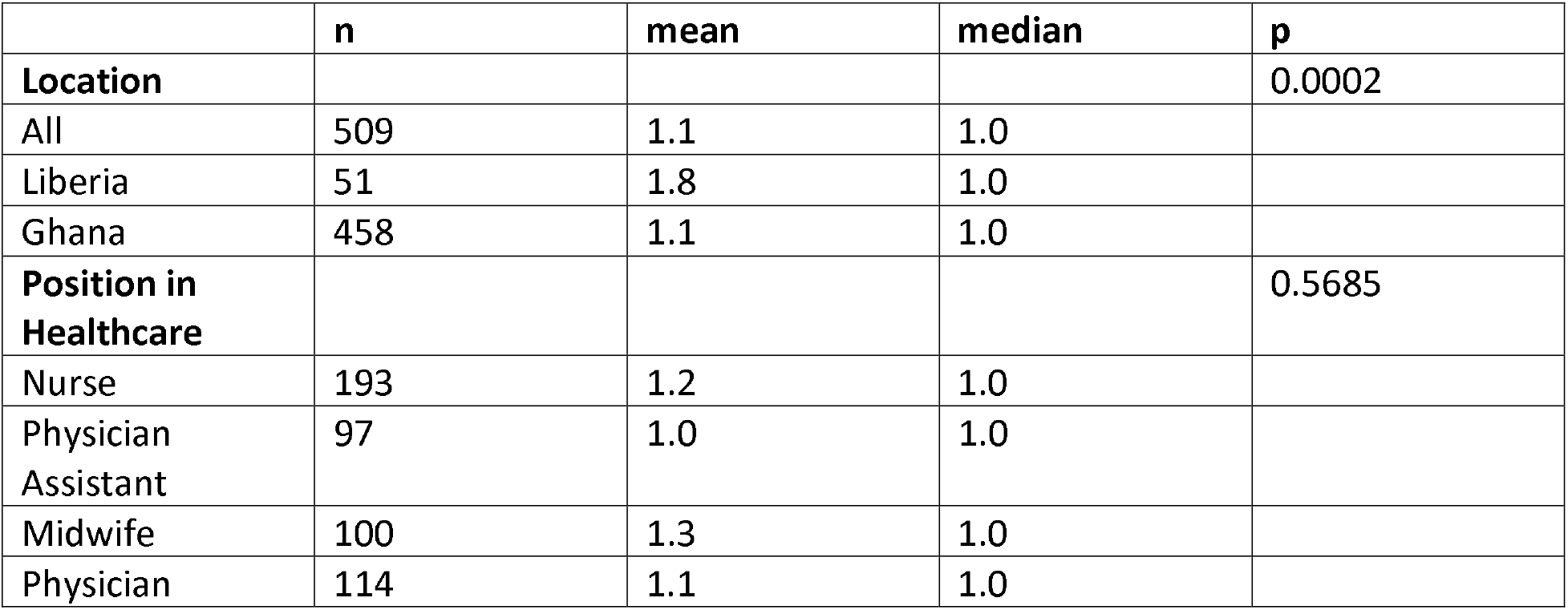
Injuries by Country and Healthcare Worker Group.

Injuries occurred in every unit type with no significant difference between the countries (*p*=0.46) (Table 3). The most frequent units with injuries were: maternity unit (20.7%), the emergency department (20.1%), and operating room (12.8%)/medical-surgical unit (12.8%). The most likely sharp to cause an injury in Ghana was a hollow needle (55.3%), followed by a solid needle (25.3%). In Liberia solid needle was likeliest to cause injury (38%), followed by a hollow needle (30.0%) (*p*<0.01).

**Table 3.** Location and Setting of Most Recent Injury.

|  | All |  | Ghana |  | Liberia |  |  |
| --- | --- | --- | --- | --- | --- | --- | --- |
|  | N | % | N | % | N | % |  |
| In which department did the needlestick occur? - Selected Choice | 492 |  | 442 |  | 50 |  | 0.4566 |
| Intensive care | 13 | 2.6% | 11 | 2.5% | 2 | 4.0% |  |
| Maternity | 102 | 20.7% | 91 | 20.6% | 11 | 22.0% |  |
| Pediatrics | 49 | 10.0% | 43 | 9.7% | 6 | 12.0% |  |
| Medical surgical | 63 | 12.8% | 60 | 13.6% | 3 | 6.0% |  |
| Injection room | 39 | 7.9% | 35 | 7.9% | 4 | 8.0% |  |
| Operating room | 63 | 12.8% | 60 | 13.6% | 3 | 6.0% |  |
| Psychiatric Unit | 5 | 1.0% | 5 | 1.1% | 0 | 0.0% |  |
| Dental Unit | 4 | 0.8% | 4 | 0.9% | 0 | 0.0% |  |
| Emergency department | 99 | 20.1% | 85 | 19.2% | 14 | 28.0% |  |
| Rotation | 13 | 2.6% | 10 | 2.3% | 3 | 6.0% |  |
| Other | 42 | 8.5% | 38 | 8.6% | 4 | 8.0% |  |
| Missing | 17 | 3.3% | 16 | 3.5% | 1 | 2.0% |  |
|  | N | % | N | % | N | % |  |
| What did you injure yourself with? | 493 |  | 443 |  | 50 |  | <0.0001 |
| Hollow needle | 260 | 52.7% | 245 | 55.3% | 15 | 30.0% |  |
| Solid bore needle | 131 | 26.6% | 112 | 25.3% | 19 | 38.0% |  |
| Scalpel | 26 | 5.3% | 18 | 4.1% | 8 | 16.0% |  |
| Don't know | 17 | 3.4% | 13 | 2.9% | 4 | 8.0% |  |
| Another sharp object | 59 | 12.0% | 55 | 12.4% | 4 | 8.0% |  |
| Missing | 16 | 3.1% | 15 | 3.3% | 1 | 2.0% |  |

Table 4 shows results for frequently injured workers, defined as workers who reported three or more injuries in the past twelve months. Overall, 15.1% of participants were in this category. Liberia had a larger proportion of such workers (29.4%) compared to Ghana (13.5%) (*p*=0.01). Across groups of healthcare workers, physician assistants had the smallest percentage of workers in this category (12.4%) while midwives had the largest (17.0%), however the difference between the two groups was not statistically significant (*p*=0.83). A higher proportion of workers who reported three or more injuries in the last year reported never receiving injury training from infection prevention and control specialists than those with fewer than three injuries (*p*=0.01).

**Table 4:** Frequently Injured Workers.

| Injuries in the last 12 months | Location |  |  |  | Position in Healthcare |  |  |  |  | P |
| --- | --- | --- | --- | --- | --- | --- | --- | --- | --- | --- |
|  | Ghana % (n) | Liberia % (n) | All % (n) | P | RN % (n) | MD % (n) | PA % (n) | RM % (n) | All % (n) |  |
| 0-2 | 86.5 (396) | 70.6 (36) | 84.9 (432) | 0.0027 | 84.5 (163) | 84.2 (96) | 87.5 (85) | 83.0 (83) | 84.7 (427) | 0.8264 |
| >=3 | 13.5 (62) | 29.4 (15) | 15.1 (77) |  | 15.5 (30) | 15.8 (18) | 12.4 (12) | 17.0 (17) | 15.3 (77) |  |
| All | 100.0 (458) | 100.0 (51) | 100.0 (509) |  | 100 (193) | 100 (114) | 100 (97) | 100 (100) | 100 (504) |  |

The overall average score on the knowledge scale, out of a possible 10, was 3.63 (Table 5). This average was calculated by adding the number of correct responses. No subtractions were made for incorrect responses. Missing responses were counted as zero. Workers who reported 0 to 2 injuries in the last 12 months had an average score of 3.69, while workers who reported 3 or more injuries had an average score of 3.30 (*p*=0.45). Further analysis indicated 54% of nurses, physicians,or physician assistants respondents who completed the scale got a score of zero, while 68% of midwives got a zero (*p*=0.09). More women (60%) than men (53%) got a zero, but this was not significant (*p*=0.126) and may be associated with midwifery, as 95% of midwives are women. Of the respondents who received a zero on the scale, 5% either didn’t know what they injured themselves with or injured themselves with another sharp object. For those participants who didn’t get a 0, that was only 1%. Removing the workers who received a score of zero on the scale resulted in a median score of 9.

**Table 5:** Knowledge and Safety Culture Scale outcomes.

|  | Knowledge Scale Scores (mean) |  | Safety Culture Scales Scores (mean) |  |
| --- | --- | --- | --- | --- |
| Overall? | 3.63 |  | 21.03 |  |
| 0-2 injuries | 3.69 | P=0.451 | 21.11 | P=0.2442 |
| >=3 injuries | 3.30 |  | 20.55 |  |

The overall average score on the CDC’s Safety Culture test, out of a possible 24, was 21. Average score on the scale was 21.1 for workers who had zero to 2 injuries in the past year and 20.6 for workers who had 3 or more (*p*=0.24).

## Discussion

This study is the first nationwide study of sharps injury incidence among healthcare workers in either Liberia or Ghana. It indicates that sharps injuries are a major occupational risk for the 142,000 healthcare workers working in those countries, with the average worker experiencing between one and two injuries per year. This is slightly lower than earlier estimates of 2.1 among healthcare workers in West Africa (7), but consistent with the estimate of 41.7% workers injured per year (2), and still indicates that these injuries are prevalent. Different worker groups reported similar frequency of injury; however, the frequency of injuries was highest in Liberia, and 66.4% of participants’ most recent injuries took place in four types of units: maternity, emergency department, operating room, and medical-surgical.

Within every healthcare worker group, between 12.4% (physician assistants) and 17.0% (midwives) of workers reported 3 or more injuries in the last year; however, the difference in percentage across groups is not statistically significant. The relatively even distribution of this frequently injured group across healthcare worker types is interesting and suggests a root vulnerability not captured in these data. A larger proportion of Liberians were a part of this frequently injured worker group.

This frequently injured population is slightly smaller than the one suggested in a previous study in Ghana (10), but significant in terms of the number of sharps injuries they experience. Further, participants in the study came solely from an emergency department (10), which we also identified as a high-risk setting. The fact that a larger percentage of these workers reported never receiving sharps training from IPC Focal People than those with fewer than three injuries suggests a training intervention may be useful to this subgroup, but further investigation into whether this subgroup has modifiable work processes or environments that increase their risk should come first. Regardless, targeting this group for intervention is promising, since reducing this group’s frequency of injury would have meaningfully lower frequency overall.

The results of the knowledge scale were intriguing. Most workers who attempted the scale received scores of zero, but among workers who did not receive a score of zero, the median knowledge score was high (9 of a possible 10). This suggests that a large proportion of healthcare workers have low sharps safety knowledge, but it is not clear what differentiates low-knowledge workers from those who scored high, as analyses on the two groups typically did not find significant differences. Midwives, the professional group with the highest average number of injuries (1.3), also had the highest percentage of participants who attempted the scale and scored zero (68%). However, neither the average number of injuries nor the rate of zero scores were significantly different for midwives than other groups.

These findings do not seem to suggest an association between low knowledge and frequent injuries, which is consistent with previous studies on occupational exposure to biological hazards in low- and middle-income countries (14). The average knowledge score of the frequently injured group barely differed from the average score of the zero to two injuries group, and the difference was not statistically significant. That said, the relative paucity of training among frequently injured workers suggests that offering training opportunities may play a valuable role beyond knowledge building, such as giving workers a chance to practice skills that require dexterity.

Overall, our findings indicate that a better understanding of the vulnerability of frequently injured workers, as associated with their work processes or environments, has the potential to be useful in lowering sharps injuries overall. A better understanding of the >50% of healthcare workers who scored zero on our knowledge scale would also be interesting, but since the association between low knowledge and injury is weak, that is a less promising avenue to approach injury reduction. The limitations of this study is the cross-sectional nature of this data limits our ability to establish causality. We cannot calculate the survey’s response rate because while the professional associations did have approximate membership numbers, they had not updated their contact information lists in some time, and it is likely some of the surveys were sent to wrong numbers. We also acknowledge the possibility that workers with recent or frequent sharps injuries were more likely to respond to this survey than workers generally. Both limitations affect generalizability.

## Data Availability

All data produced in the present study are available upon reasonable request to the authors, as is the survey instrument.

## Funding Sources for ALL Authors

This study was funded by the University of Michigan’s Center of Occupational and Safety Engineering. The PI was also funded by as a T32 fellow in Complexity: Innovations in Promoting Health and Safety (CIPHS), National Institute of Nursing Research (T32 NR016914, PI: Titler) at the time of data collection. This study did not obtain open access funding from either source.

## Conflict of Interest

NONE DECLARED

## Acknowledgments

The authors would like to acknowledge the work of Mr. Anthony Arkoh and the Graduate Physician Assistants Association of Ghana, Ms. Frederica Hanson and the Ghana Registered Midwives Association, Dr. Kofi Mensah Boateng and the Junior Doctors’ Association of Ghana, Mr.Benjamin Suamey and the Liberian Nurses Association, Ms. Shirley Fahnbulleh and the West African College of Nurses, Ms. Wilhelmina Flomo and the Liberian Midwives Association, and Mr. Theophilus Fayiah and the Physician Assistant Association of Liberia.

## Notes

### Competing Interest Statement

The authors have declared no competing interest.

### Funding Statement

This study was funded by the University of Michigan Center of Occupational and Safety Engineering. The PI was also funded by as a T32 fellow in Complexity: Innovations in Promoting Health and Safety (CIPHS), National Institute of Nursing Research (T32 NR016914, PI: Titler) at the time of data collection.

### Author Declarations

This project received in-country approval from the University of Liberia Institutional Review Board (Protocol #21-12-296) and the Ghana Health Services Ethics Research Committee (GHS ERC Number: 018/11/2021). It was deemed exempt by the Institutional Review Board of the University of Michigan and the University of Cincinnati.

